# Elevated levels of the cytokine LIGHT in Crohn’s disease

**DOI:** 10.1101/2021.07.16.21260656

**Authors:** Christopher J. Cardinale, Debra J. Abrams, Frank D. Mentch, John A. Cardinale, Charlly Kao, Patrick M.A. Sleiman, Hakon Hakonarson

**Author notes:** Corresponding Author: Hakon Hakonarson, MD, PhD, Center for Applied Genomics, Children’s Hospital of Philadelphia, Abramson Bldg Ste 1216, 3615 Civic Center Blvd, Philadelphia, PA 19104.

## Abstract

LIGHT, encoded by the *TNFSF14* gene, is a cytokine belonging to the tumor necrosis factor superfamily. Upon binding to its receptors, HVEM and LTBR, it activates inflammatory responses. We used a single-molecule immunoassay to determine the circulating levels of free LIGHT in plasma from pediatric patients with Crohn’s disease (N = 183) and a panel of healthy pediatric reference samples (N = 9). LIGHT levels were greatly elevated in Crohn’s disease (average of 305 pg/ml *versus* 57 pg/ml in controls, *P* < 0.0001). We performed correlational analyses between LIGHT levels and the clinical characteristics of the Crohn’s cohort, including age, Montreal classification, family history, medical/surgical therapy, and routine blood test parameters. We found statistically significant correlation between white blood cell count and free LIGHT (*P* < 0.046). Our results support the hypothesis that elevated levels of the cytokine contribute to the pathology of Crohn’s disease and that therapies to neutralize free LIGHT with antibodies may be beneficial.

## Introduction

Crohn’s disease is a condition of the gastrointestinal tract in which the immune system undergoes an inappropriate and destructive inflammatory reaction against normal gut contents, such as food and commensal microorganisms (1). In contrast to the other major type of inflammatory bowel disease (IBD), ulcerative colitis, Crohn’s lesions can occur at any point in the digestive system from mouth to anus, although the terminal ileum of the small bowel and the colon are most frequently affected. Worrisome complications of Crohn’s disease include a deep, full-thickness inflammation of the bowel, with possible stricturing or fistulization, as well as lesions around the anus which can be painful and infected. In addition to frequent diarrhea and abdominal pain, malabsorption can result in delayed growth and development in the pediatric population (2).

The mainstay of treatment for Crohn’s disease is immunosuppression, frequently with neutralizing monoclonal antibodies to tumor necrosis factor, such as infliximab or adalimumab. Other biologics in use include ustekinumab, a blocker of IL-12 and IL-23, and vedolizumab, an antibody to integrin α_4_β_7_. Non-biologic drugs include salicylates (e.g., mesalamine), thiopurines (e.g., azathioprine), and steroids (e.g., budesonide), all of which act to reduce inflammation. Frequently, surgery is required to resect ileum or other lesions in the GI tract. Despite these treatments, Crohn’s disease tends to progress over the lifespan, with significant impact on health and quality of life. Therefore, new treatments are seriously needed.

In our efforts to develop new therapies, our attention was drawn to a tumor necrosis factor superfamily cytokine known as LIGHT, or HVEM-L (*TNFSF14*). LIGHT (homologous to *l*ymphotoxins, exhibits *i*nducible expression, and competes with HSV *g*lycoprotein D for *H*VEM, a receptor expressed by *T* lymphocytes) activates TNF receptor superfamily members herpes virus entry mediator (HVEM, *TNFRSF14*) and lymphotoxin β receptor (*LTBR*), which are expressed on T cells. LIGHT is inactivated by Decoy Receptor 3 (*TNFRSF6B*), a secreted, non-signaling TNF superfamily receptor. This network of ligands and receptors regulates innate and adaptive immune responses systemically, as well as in the mucosal immune system. Our interest in this network was aroused by the very strong associations observed in genome-wide association studies (GWAS) of Crohn’s and inflammatory bowel diseases, in which the TNF superfamily genes played a leading role (3, 4).

The numerous genetic signals for this network in IBD suggest that a monoclonal antibody which inhibits LIGHT’s signaling to HVEM on T cells would have beneficial immunomodulatory effects. We therefore sought to establish whether circulating levels of free LIGHT are elevated in Crohn’s disease patients. The cytokine is free in the sense that its transmembrane domain has been cleaved, resulting in a soluble ligand. We expect that this free level is a proxy of LIGHT’s ability to stimulate T cell responses. Our results show that pediatric Crohn’s disease patients have LIGHT levels that are 5-fold higher than healthy controls. The increased LIGHT level is consistent across different disease activities, locations, and therapies.

## Results

We identified a group of 183 Crohn’s disease patients with available plasma samples from the general-purpose pediatric biobank of the Center for Applied Genomics of the Children’s Hospital of Philadelphia, consisting of over 80,000 enrolled subjects. All patients have been diagnosed and treated at this hospital with regular follow-up. Summary statistics on the clinical makeup of the cohort are given in Table 1. This population has high rates of surgery and biologic therapy use, as would be expected for a tertiary referral hospital with a specialized program in IBD. The cases are 80% Caucasian with approximately equal male and female subjects. Plasma samples were collected between 2006 and 2018 and remained in frozen storage. These samples were tested along with nine purchased reference controls of healthy pediatric plasma at a services lab using the Simoa single-molecule immunoassay (Quanterix). The Crohn’s disease patients exhibited an average free LIGHT level of 305 pg/ml, in comparison to the reference samples, which averaged 57 pg/ml (*P* < 0.0001) (Fig. 1). In this sample, 83% of Crohn’s disease subjects had LIGHT levels exceeding two standard deviations above the average of the controls (> 128 pg/ml).

**Table 1.** Summary statistics of the Crohn’s disease cohort, overall N = 183.

| **Trait** | **Cases (SD)** | **Range** | **Controls (SD)** |
| --- | --- | --- | --- |
| Age at sample collection (y) | 14.6 (4.0) | 1.2–22.8 | 13.3 (1.9) |
| Age at Diagnosis (y) | 11.4 (3.8) | 0.5–18.0 |  |
| Sex (% male) | 57% |  | 67% |
| Family history (1<sup>st</sup>-degree) | 20% |  |  |
| **Race** |  |  |  |
| White/Caucasian | 80.3% |  | 67% |
| African-American | 12.6% |  | 33% |
| Mixed or non-White/non-African | 7.1% |  | 0% |
| **Location (Montreal)** |  |  |  |
| Upper GI involvement | 74% |  |  |
| Ileal-only | 17% |  |  |
| Ileocolonic | 73% |  |  |
| Colonic-only | 12% |  |  |
| **Behavior (Montreal)** |  |  |  |
| Non-stricturing, non-penetrating | 53% |  |  |
| Stricturing | 33% |  |  |
| Penetrating | 28% |  |  |
| Perianal involvement | 27% |  |  |
| **Treatment** |  |  |  |
| Surgery | 31% |  |  |
| Biologic drug | 67% |  |  |
| **LIGHT level (pg/ml)** | 305 (210) | 31–1080 | 57.1 (35.4) |

**Figure 1.**
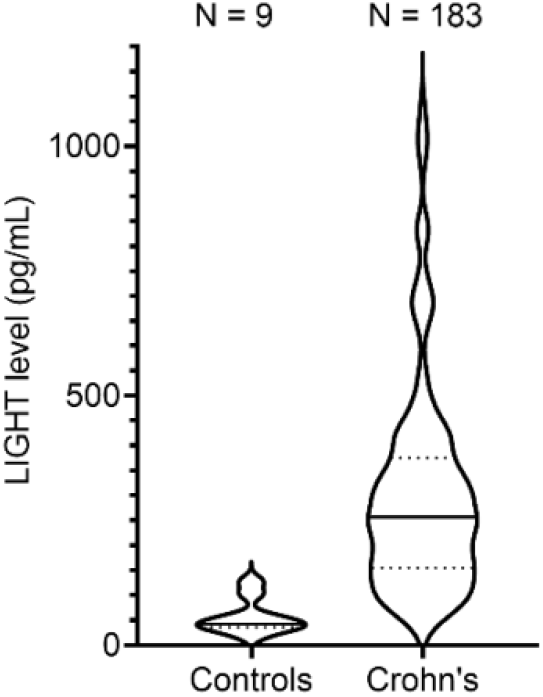
Levels of free LIGHT cytokine are elevated in pediatric Crohn’s disease patients compared to a panel of healthy reference samples. Levels were determined by Simoa single-molecule immunoassay on preserved plasma samples from a hospital research biobank and displayed as a violin plot. Horizontal lines in the “violins” indicate the 25^th^, 50^th^, and 75^th^ percentiles. Mann-Whitney U test show a statistically significant difference between the controls and Crohn’s patients (*P* < 0.0001).

We investigated the relationship between clinical characteristics of the patients and the level of free LIGHT. These sub-phenotypes included age, sex, disease behavior and location according to the Montreal classification, family history, and history of surgery or biologic therapy. The only non-zero correlation was found with the white blood cell count, showing a modest increase in LIGHT levels associated with higher white counts (*P* < 0.046). These observations demonstrate that Crohn’s disease patients exhibit highly elevated LIGHT levels, but that specific disease characteristics are not systematically associated (Fig. 2). This result is unsurprising considering the cross-sectional nature of the study, in which samples were collected independent of the current disease activity, duration of illness, or treatment with anti-inflammatory agents.

**Figure 2.**
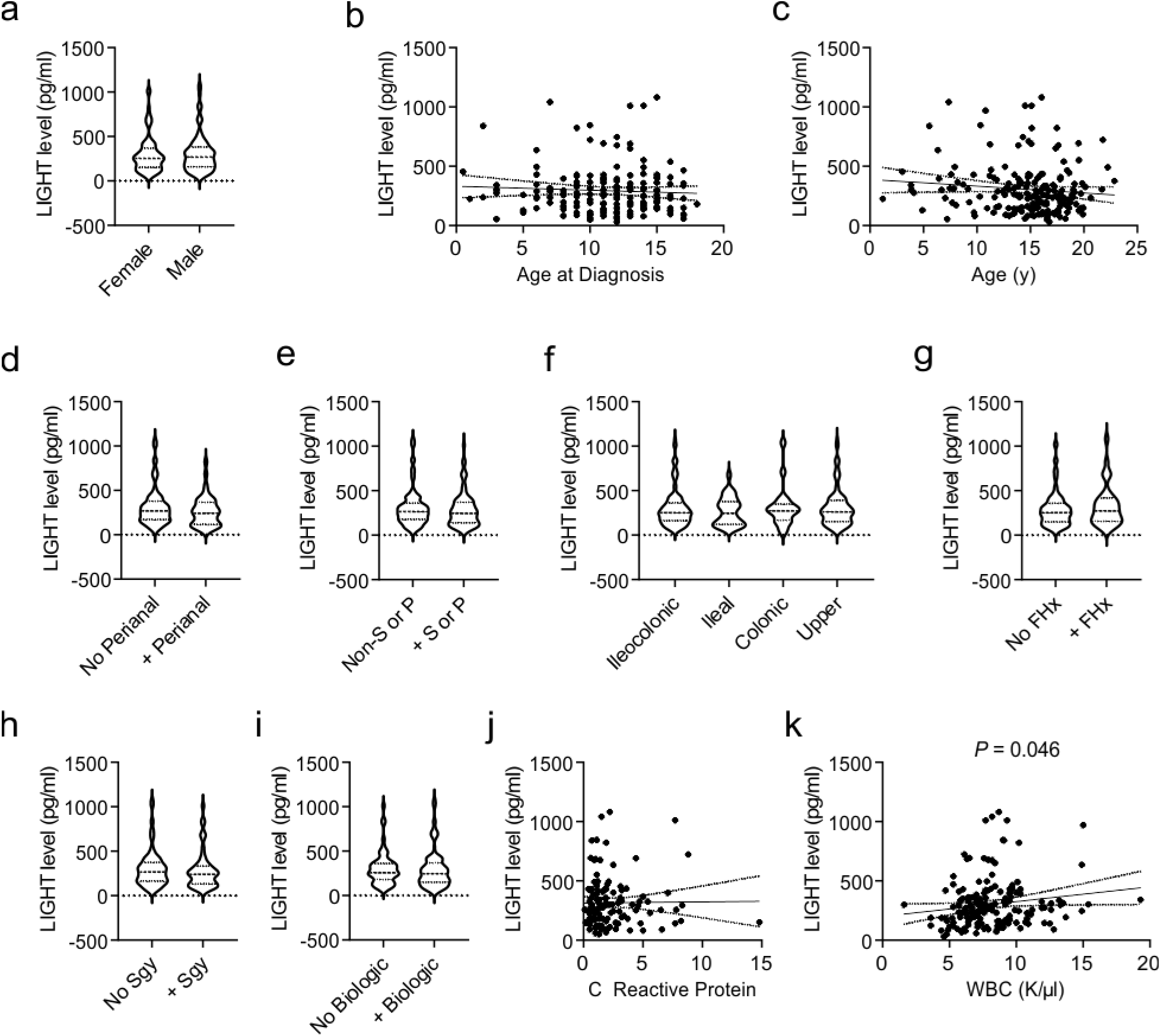
Plasma free LIGHT levels are stable in Crohn’s disease patients across a wide range of phenotypes. Violin plots are displayed of the LIGHT level in different clinical parameters including (a) sex, (b) age at diagnosis, (c) age of subject at sample collection, (d) presence of perianal involvement, (e) presence of structuring or penetrating disease behavior, (f) disease location, (g) presence of family history in a 1^st^-degree relative, (h) history of surgery for Crohn’s disease, (i) history of biologic therapy with infliximab or adalimumab, (j) level of C Reactive Protein within 6 months of sample collection, and (k) white blood cell count on hemogram. Only the slope of the regression line of WBC *versus* LIGHT was significantly non-zero (*P* = 0.046 by *F*-test).

## Discussion

The presence of high levels of free LIGHT, combined with genetic associations between polymorphisms in the TNF superfamily genes and disease status, argues that this cytokine plays an intimate role in the pathogenesis of Crohn’s disease. Future studies should include prospective trials in which subjects are tested according to specific criteria of current disease activity and sampled longitudinally. Hypothetically, LIGHT levels could be elevated during acute flares, when much of the tissue damage occurs.

In the TNBS and dextran sulfate models of colitis in mice, neutralization of LIGHT reduced inflammation and promoted healing (5, 6). Transgenic mice expressing LIGHT on T cells had increased intestinal inflammation (7), while conversely, mice lacking LIGHT also show greater susceptibility to colitis (8, 9). The explanation for these apparently discordant results lies in the complex interplay of different combinations of ligands and receptors in the TNF family network.

Clinical trials are currently underway to determine if CERC-002, a monoclonal antibody that neutralizes LIGHT, can improve disease outcomes in Crohn’s disease. This antibody could make a useful addition to the armamentarium of biologic treatments available to TNF-non-responders. Primary non-response to anti-TNF therapy is found in 30–40% of Crohn’s disease patients, and further individuals may lose response (10). Furthermore, primary non-responders are more likely to fail second-line agents (11). Therefore, exploration of alternative treatments is called for.

## Materials and Methods

### Design

Reference samples of pediatric plasma were obtained from BioIVT (Westbury, N.Y.) to match the age, sex, and race composition of the Crohn’s disease cohort. The patients were recruited through the Center for Applied Genomics as part of its pediatric biobank program. The patients were diagnosed by pediatric gastroenterologists at the Children’s Hospital of Philadelphia using standard criteria (12), including physical symptoms, radiologic studies, endoscopy, and tissue biopsies. The cohort was assembled from the biobank using searches of the de-identified electronic medical record (Epic) and research intake survey to select individuals with frozen plasma samples available.

In addition to queries of discrete fields in the Epic record, a genetic counselor (D.J.A.) reviewed clinical notes including gastroenterology visits, discharge summaries, and biopsy/endoscopy reports to ascertain the subphenotypes of the disease cohort.

### LIGHT immunoassay

Free LIGHT was determined using the single-molecule array immunoassay (Simoa) technology from Quanterix. The assays were performed in an outside services lab (Myriad RBM, Austin, Tx.). Briefly, LIGHT was captured by antibody immobilized on magnetic beads followed by a biotinylated detection antibody. The beads were washed and incubated with streptavidin-β-galactosidase. The beads were loaded on the Simoa disc, incubated with the fluorescent enzyme substrate, and scanned. In this method, the number of positive beads, rather than the intensity of the fluorescence, is used to determine the level of free LIGHT compared with recombinant standards, resulting in high sensitivity and wide dynamic range (0.8 to 4000 pg/ml).

The plasma samples were collected between 2006 and 2018 and were kept at −80°C. The levels of free LIGHT remained stable over this period of storage (Fig. 3).

**Figure 3.**
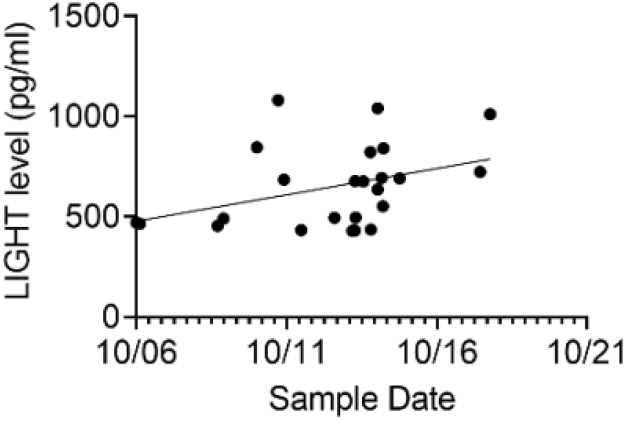
LIGHT levels show a modest decline in older samples, as shown by the collection dates of the 25 highest levels measured in the cohort.

### Statistics

The clinical characteristics and LIGHT levels of the Crohn’s disease cohort was tabulated in a spreadsheet and in Prism 9 (GraphPad Software). The comparison between cases and controls was by Mann-Whitney U test (in Prism), because the LIGHT levels are not normally distributed. For continuous traits (white blood cell levels, age at sample collection, etc) a simple linear regression was computed with an *F*-test to determine if the slope is significantly non-zero. For categorical variables (biologic therapy, perianal involvement, etc) a Mann-Whitney U test was used to compare groups.

## Data Availability

Data is available from the corresponding author on request.

## Acknowledgment

Funding was provided by a special purpose fund from Cerecor, Inc.

